# Melatonin is significantly associated with survival of intubated COVID-19 patients

**DOI:** 10.1101/2020.10.15.20213546

**Authors:** Vijendra Ramlall, Jason Zucker, Nicholas Tatonetti

## Abstract

**Background:** Respiratory distress requiring intubation is the most serious complication associated with coronavirus disease 2019 (COVID-19).

**Methods:** In this retrospective study, we used survival analysis to determine whether or not mortality following intubation was associated with hormone exposure in patients treated at New York Presbyterian/ Columbia University Irving Medical Center. Here, we report the overall hazards ratio for each hormone for exposure before and after intubation for intubated and mechanically ventilated patients.

**Results:** Among the 189,987 patients, we identified 948 intubation periods across 791 patients who were diagnosed with COVID-19 or infected with SARS-CoV2 and 3,497 intubation periods across 2,981 patients who were not. Melatonin exposure after intubation was statistically associated with a positive outcome in COVID-19 (demographics and comorbidities adjusted HR: 0.131, 95% CI: 7.76E-02 - 0.223, p-value = 8.19E-14) and non-COVID-19 (demographics and comorbidities adjusted HR: 0.278, 95% CI: 0.142 - 0.542, p-value = 1.72E-04) intubated patients. Additionally, melatonin exposure after intubation was statically associated with a positive outcome in COVID-19 patients (demographics and comorbidities adjusted HR: 0.127, 95% CI: 6.01E-02 - 0.269, p-value = 7.15E-08).

**Conclusions:** Melatonin exposure after intubation is significantly associated with a positive outcome in COVID-19 and non-COVID-19 patients. Additionally, melatonin exposure after intubation is significantly associated with a positive outcome in COVID-19 patients requiring mechanical ventilation. While our models account for many covariates, including clinical history and demographics, it is impossible to rule out confounding or collider biases within our population. Further study into the possible mechanism of this observation is warranted.

## Introduction

The coronavirus disease 2019 (COVID-19) pandemic caused by severe acute respiratory syndrome coronavirus 2 (SARS-CoV2) infection has impacted every country in the world with more than 35 million confirmed cases and more than 1 million deaths globally - the United States accounts for more than 20% of total cases and deaths[1]. In the months since the first infections were reported outside of the original epicenter, clinical research remains focused on identifying treatments[2,3,4] and preventive measures[5,6,7] for SARS-CoV2 infection.

Analyses of healthcare data from infected patients have identified the most frequent symptoms, e.g. fever, cough, fatigue, shortness of breath, loss of taste or smell[8], however less frequent symptoms influenced by comorbidities are also observed[9,10,11]. Respiratory distress remains the most significant and serious symptom of COVID-19[12], which, in the most severe of cases, can require endotracheal intubation and mechanical ventilation, and even lung transplants[13]. At the core of the public health emergency that has ravaged the world, is the limited amount of supplies and number of intensive care unit beds[14]. Furthermore, patients requiring respiratory support, intubation, oxygen supplementation or invasive mechanical ventilation are bearing the brunt of the limited availability of resources[14].

Among the possible therapies for SARS-CoV2 infection being researched, hormone drugs, such as dexamethasone[15] and methylprednisolone[16], have proved promising. The dexamethasone study from the RECOVERY Collaborative Group in the UK found that patients overall (22.9%), patients requiring invasive mechanical ventilation (29.3%) and patients receiving oxygen without mechanical ventilation (23.3%) treated with dexamethasone had lower death rates at 28 days compared to those who were treated with usual care (25.7%, 41.4% and 26.2%, respectively)[15].

Here, we present a retrospective study on the effects of hormone exposure in COVID-19 patients at New York Presbyterian/ Columbia University Irving Medical Center (NYP/CUIMC), who required endotracheal intubation, on mortality. Our univariate analysis indicated that melatonin exposure after intubation was significantly associated with survival in intubated COVID-19 patients (HR: 9.17E-02, 95% CI: 5.43E-03 - 0.155, p-value = 4.81E-19) and mechanically ventilated COVID-19 patients (HR: 9.13E-02, 95% CI: 4.40E-02 - 0.189, p-value = 1.32E-10). Our multivariate analysis accounting for significant demographic and clinical diagnoses indicated that melatonin exposure was significantly associated with survival in COVID-19 patients (HR: 0.131, 95% CI: 7.66E-02 - 0.223, p-value = 8.19E-14) and in mechanically ventilated COVID-19 patients (HR: 0.127, 95% CI: 6.01E-02 - 0.269, p-value = 7.15E-08). Moreover, our univariate analysis indicated that treatment with dexamethasone after intubation was associated, though not significantly, with a positive outcome in intubated COVID-19 patients (HR:0.167, 95% CI: 2.35E-02 - 1.19, p-value =7.43E-2). While our analysis identified significant associations between melatonin exposure after intubation and survival, we have not identified the directionality nor the underlying mechanism nor can we rule out collider bias from our analysis.

## Results

### Identifying patient cohorts

We conducted a retrospective observational study of 189,987 patients who sought care at NYP/ CUIMC between February 1st, 2020 and August 1st, 2020. We identified 13,394 patients who were diagnosed with COVID-19 or infected with SARS-CoV2 and 948 intubation periods among the 791 patients who required endotracheal intubation. Additionally, we identified 3,497 intubation periods among the 2,981 patients who required endotracheal intubation and were not diagnosed with COVID-19 nor infected with SARS-CoV2, which served as the controls in this study (Table 1). From our clinical data warehouse (CDW), we identified 747 intubation periods among the 637 patients who required endotracheal intubation between February 1st, 2018 and August 1st, 2018 (Table 1). Of the intubation periods for COVID-19 patients, 315 required mechanical ventilation and 276 resulted in death within seven days of extubation (i.e. negative outcome), 242 non-COVID-19 intubation periods required mechanical ventilation and 143 resulted in death within seven days of extubation, 637 intubation periods from 2018 required mechanical ventilation and 174 resulted in death within seven days of extubation (Table 1).

**Table 1.** Frequency of demographics, diseases and outcome of intubation periods’ patients

|  | **COVID19 +** | **COVID19 -** | **2018** |
| --- | --- | --- | --- |
| **N** | 948 | 3497 | 747 |
| **Average Age (IQR)** | 56.52<br>(0.0, 95.14) | 46.44<br>(0.0, 97.72) | 52.62<br>(0.0, 118.00) |
| **Age ≥ 65** | 409<br>43.14% | 1125<br>32.17% | 315<br>42.17% |
| **Male** | 577<br>60.86% | 1774<br>50.73% | 440<br>58.90% |
| **Race and Ethnicity** |  |  |  |
| **Hispanic or Latin or Spanish origin** | 438<br>46.20% | 898<br>25.68% | 151<br>20.21% |
| **American Indian or Alaskan Native** | ≤10<br>≤1.05% | 11<br>0.31% | ≤10<br>≤1.34% |
| **Asian** | 13<br>1.37% | 76<br>2.17% | 11<br>1.47% |
| **Black or African American** | 195<br>20.57% | 548<br>15.67% | 112<br>14.99% |
| **Native Hawaiian or Other Pacific Islander** | ≤10<br>≤1.05% | ≤10<br>≤0.27% | ≤10<br>≤1.34% |
| **White** | 269<br>28.38% | 1551<br>44.35% | 267<br>35.74% |
| **Disease** |  |  |  |
| **Asthma** | 135<br>14.24% | 336<br>9.61% | 148<br>19.81% |
| **Cardiovascular disease** | 662<br>69.83% | 2149<br>61.45% | 723<br>96.79% |
| **Chronic kidney disease** | 671<br>70.78% | 1303<br>37.26% | 259<br>34.67% |
| **Chronic obstructive lung disease** | 85<br>8.97% | 244<br>6.98% | 187<br>25.03% |
| **Coronary artery disease** | 145<br>15.30% | 539<br>15.41% | 265<br>35.48% |
| **Diabetes mellitus** | 408<br>43.04% | 722<br>20.65% | 283<br>37.88% |
| **Diabetic nephropathy** | 56<br>5.91% | 107<br>3.06% | 64<br>8.57% |
| **Diabetic neuropathy** | 52<br>5.49% | 104<br>2.97% | 57<br>7.63% |
| **Diabetic retinopathy** | 28<br>2.95% | 48<br>1.37% | 23<br>3.08% |

Table 1 Frequency of demographics, diseases and outcome of intubation periods' patients, cont.
|  | COVID19 + | COVID19 - | 2018 |
| --- | --- | --- | --- |
| Disease |  |  |  |
| Diabetic vasculopathy | 26<br>2.74% | 58<br>1.66% | 38<br>5.09% |
| Heart failure | 198<br>20.89% | 518<br>14.81% | 387<br>51.81% |
| Hypertension | 415<br>43.78% | 1181<br>33.77% | 471<br>63.05% |
| Myocardial infarction | 80<br>8.44% | 216<br>6.18% | 160<br>21.42% |
| Obesity | 450<br>47.47% | 1225<br>35.03% | 193<br>25.84% |
| Respiratory disease | 706<br>74.47% | 1819<br>52.02% | 746<br>99.87% |
| Outcome |  |  |  |
| Mechanical ventilation required | 315<br>33.23% | 242<br>6.92% | 637<br>85.27% |
| Death within 7 days of extubation | 276<br>29.11% | 143<br>4.09% | 174<br>23.29% |

The median (interquartile range) age of the COVID-19, non-COVID-19, and 2018 patients with intubation periods was 56.52 (0 - 95.14) years, 46.44 (0-97.72) years and 52.62 (0.-118.00), respectively, and 60.86%, 50.73% and 58.90%, respectively, self-identified as male (Table 1). Additionally, 1.37%, 20.57% and 28.38% of the COVID-19 intubation periods, 2.17%, 5.67% and 44.35% of non-COVID-19 intubation periods and 1.47%, 14.99%, 35.74% of intubation periods in 2018 were for patients who self-identified as Asian, Black or African American and White, respectively, and 46.20% of the COVID-19 patients for COVID-19 patients who identified as of Hispanic or Latin or Spanish origin compared to 25.68% of the non-COVID-19 intubations and 20.21% of intubation periods from 2018 (Table 1). More than 50% of the COVID-19, non-COVID-19 and 2018 intubation periods were for patients who had a history of cardiovascular disease and respiratory disease (Table 1). Additionally, 70.78%, 43.04%, 20.89%, 43.78% and 47.47% of the COVID-19 intubations periods were for patients who had a history of chronic kidney disease, diabetes mellitus, heart failure, hypertension and obesity, respectively, compared to 37.36%, 20.65%, 14.81%, 33.77% and 35.05% of the non-COVID-19 intubation periods and 34.67%, 37.88%, 51.81%, 63.05% and 25.84% of intubation periods from 2018 (Table 1).

### Univariate analysis of hormone exposure on outcome following intubation

We used univariate analysis of hormone exposures following intubation to identify hypotheses for follow up analysis. Among the subset of intubation periods for COVID-19 patients during which the patient required mechanical ventilation, exposure to methylprednisolone (HR: 1.63, 95% CI: 1.07 - 2.47 p-value = 2.37E-02) and levothyroxine (HR: 2.26, 95% CI: 1.13 - 4.51, p-value = 2.04E-02) were significantly associated with a negative outcome (Table S3, Figure S1).

Exposure to insulin glargine (HR: 0.665, 95% CI: 0.521 - 0.849 p-value = 1.04E-03), budesonide (HR: 0.290, 95% CI: 0.108 - 0.778, p value = 1.40E-02), melatonin (HR: 9.17E-02, 95% CI: 5.43E-02 - 0.155, p-value = 4.81E-19), prednisone (HR: 0.432, 95% CI: 0.230 - 0.812. p-value = 9.11E-03), methylprednisolone (HR: 0.773, 95% CI: 0.603 - 0.991, p-value = 4.25 E-02) and insulin lispro (HR: 0.731, 95% CI: 0.575 - 0.930, p-value = 1.07E-02) between the intubation day and the extubation day were significantly associated with a positive outcome in intubation periods of COVID-19 patients (Table S3, Figure S1). Among the same intubation periods, exposure to hydrocortisone (HR: 1.56, 95% CI: 1.22 - 2.00, p-value = 3.54E-04) between the intubation day and the extubation day was significantly associated with a negative outcomes (Table S3, Figure S1).

Exposure to melatonin (HR: 9.13E-02, 95% CI: 4.40E-02 - 0.189, p-value = 1.32E-10) between the intubation day and the extubation day was significantly associated with a positive outcome in intubation periods for COVID-19 patients requiring mechanical ventilation (Table S3, Figure S2). Conversely, exposure to hydrocortisone (HR: 2.16 95% CI: 1.42 - 3.28, p-value = 2.98E-04), methylprednisolone (HR: 1.73, 95% CI: 1.13 - 2.64, p-value = 1.19E-02) and levothyroxine (HR: 1.89, 95% CI: 1.05 - 3.40, p-value = 3.43E-02) between the intubation day and the extubation day was significantly associated with a negative outcome (Table S3, Figure S2).

### Dexamethasone treatment after intubation is associated with increased survival among intubated COVID-19 patients

Based on the results of the study from the RECOVERY Collaborative Group in the UK, we were interested in the effects of dexamethasone treatment when accounting for other significant covariates, following the univariate analysis. We fit a Cox proportional hazards model using age (binary for whether the patients was older than 65), whether or not the patient self-identified as American Indian or Alaskan Native, whether or not the patient had a history of chronic kidney disease, chronic obstructive lung disease, coronary artery disease, diabetes mellitus, hypertension, myocardial infarction and respiratory disease, and whether or not a patient was exposed to dexamethasone after being intubated. Being at least 65 years old (HR: 2.45, 95% CI: 1.88 - 3.20, p-value = 4.14E-11) and having a history of chronic kidney disease (HR: 4.05, 95% CI: 2.28 - 7.18, p-value = 1.82E-06) and a history of chronic obstructive lung disease (HR: 1.48, 95% CI: 1.01 - 2.17, p-value = 4.25E-02) were significantly associated with a negative outcome following intubation in intubation periods for COVID-19 patients (Table S4). Having a history of respiratory disease (HR: 0.470, 95% CI: 0.351 - 0.631, p-value = 4.71E-07) was significantly associated with a positive outcome in intubation periods for COVID-19 patients (Table S4). While not significant in the model, exposure to dexamethasone after intubation was associated with a positive outcome (HR: 0.235, 95% CI: 3.25E-02 - 1.69, p-value = 0.151) in intubation for COVID-19 patients (Table S4).

In similar model, being at least 65 years old (HR: 1.93, 95% CI: 1.3 4-2.78, p-value = 3.84E-04), having a history of chronic kidney disease (HR: 4.45, 95% CI: 2.80 - 7.09, p-value = 3.12E-10) and a history of myocardial infarction (HR: 2.55, 95% CI: 1.59 - 4.09, p-value = 1.04E-04) were significantly associated with a negative outcome in intubation periods for non-COVID-19 patients (Table S4). Being at least 65 years old (HR: 2.48, 95% CI: 1.71 - 3.58, p-value = 1.41E-06) and having a history of chronic kidney disease (HR: 1.44, 95% CI: 1.03 - 2.01, p-value = 3.16E-02) were significantly associated with a negative outcome in intubation periods from 2018 (Table S4).

In a follow up analysis of the intubation periods where the patient required mechanical ventilation, we fit a Cox proportional hazards model using age (binary for whether the patients was older than 65), whether or not the patient self-identified as Black or African American, whether or not the patient had a history of asthma, chronic kidney disease, chronic obstructive lung disease and respiratory disease and whether or not a patients was exposed to dexamethasone after intubation. Being at least 65 years old (HR: 2.29, 95% CI: 1.46 - 3.59, p-value = 3.00E-04) was significantly associated with a negative outcome in intubation periods for COVID-19 patients (Table S5). Self-identifying as Black or African American (HR: 0.384, 95% CI: 0.185 - 0.797, p-value = 1.02E-02) and having a history of respiratory disease (HR: 0.534, 95% CI: 0.314 - 0.910, p-value = 2.11E-02) were significantly associated with a positive outcome in intubation periods for COVID-19 patients (Table S5).

In a similar model, being at least 65 years old (HR: 3.11, 95% CI: 1.47 - 6.55, p-value = 2.91E-03) was associated with a negative outcome in intubation periods for non-COVID-19 patients (Table S5). Being at least 65 years old (HR: 2.77, 95% CI: 1.90 - 4.03, p-value = 1.11E-07) was associated with a negative outcome in intubation periods from 2018 (Table S5).

### Univariate analysis of melatonin, quetiapine, trazodone and benzodiazepines on outcome following intubation

Following the significant association between melatonin exposure following intubation and a positive outcome in intubation periods and in intubation periods requiring mechanical ventilation for COVID-19 patients, we conducted a univariate analysis of exposure to quetiapine, trazodone and benzodiazepines in COVID-19 and non-COVID-19 patients. Exposure to quetiapine (HR: 0.536, 95% CI: 0.293 - 0.980, p-value = 4.28E-02) and benzodiazepines (HR: 0.477, 95% CI: 0.330 - 0.690, p-value = 8.31E-05) between the visit start day and the intubation day were significantly associated with a positive outcome following intubation in intubation periods for COVID-19 patients (Table S6, Figure 3). Exposure to quetiapine (HR: 0.242, 95% CI: 0.178 - 0.329, p-value = 1.60E-19), trazodone (HR: 8.31E-02, 95% CI: 1.17E-02 - 0.593, p-value = 1.13E-02) and benzodiazepines (HR: 0.418. 95% CI: 0.318 - 0.550, p-value = 4.18E-10) between the intubation day and the extubation day were significantly associated with a positive outcome in intubation periods for COVID-19 patients (Table S6, Figure 2). Among intubation periods for non-COVID-19 patients, exposure to benzodiazepines (HR: 0.322. 95% CI: 0.232 - 0.447, p-value = 1.48E-11 between the intubation day and the extubation day were significantly associated with a positive outcome for intubation for non-COVID-19 patients (Table S6, Figure 2). Exposure to quetiapine (HR: 0.471, 95% CI: 0.328 - 0.676, p-value = 4.46E-05) was significantly associated with a positive outcome in intubation periods from 2018 (Table S6).

**Figure 1.**
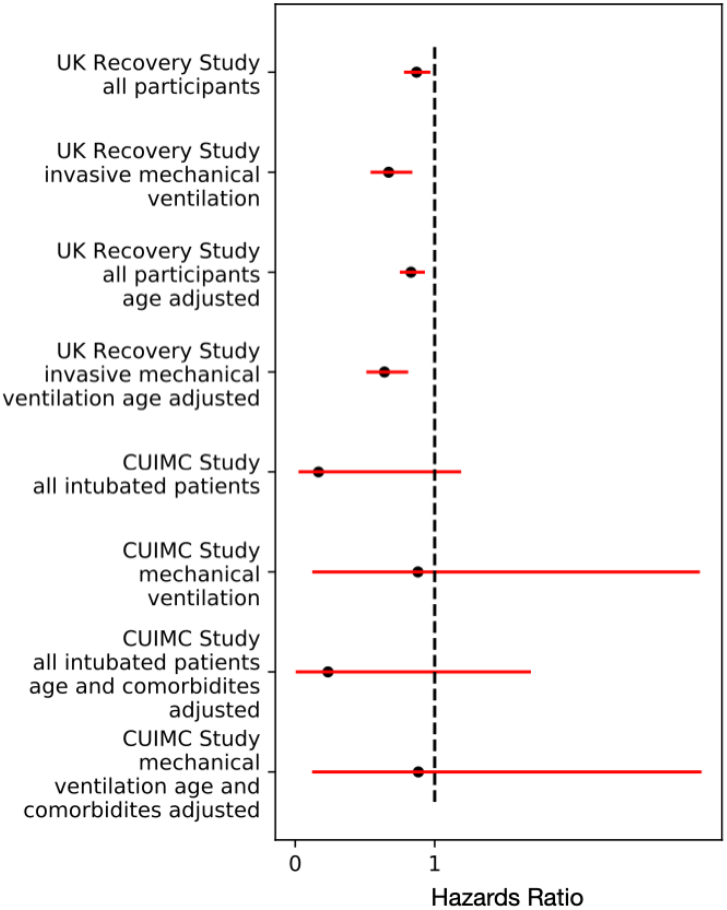
Comparison of dexamethasone export hazard ratios from the RECOVERY Study and this study for COVID-19 patients.

**Figure 2.**
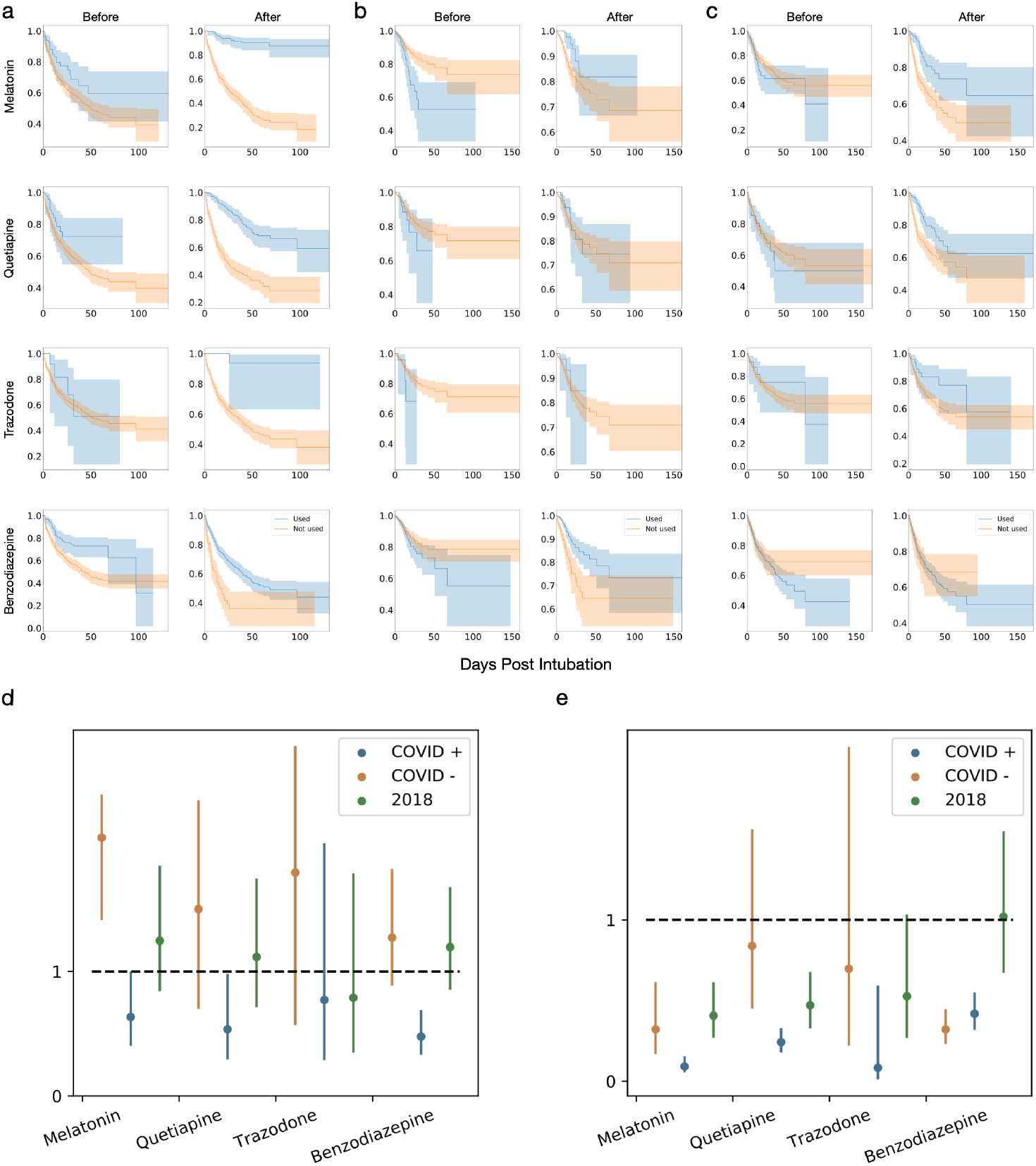
a-c Kaplan-Meier curves for melatonin, quetiapine, trazodone and benzodiazepines treatment before and after intubation for (a) COVID-19 intubation periods, (b) non-COVID-19 intubation periods and (c) intubation periods from 2018. d,e. Comparison of hazard ratios for melatonin, quetiapine, trazodone and benzodiazepines exposure (d) before and (e) after intubation.

### Melatonin treatment is associated with increased survival among intubated patients

In order to further understand the association between melatonin exposure after intubation and survival, we were interested in the effect of melatonin exposure when accounting for other factors significant covariates. We fit a Cox proportional hazards model using age (binary for whether the patients was older than 65), whether or not the patient self-identified as American Indian or Alaskan Native, whether or not the patient had a history of chronic kidney disease, chronic obstructive lung disease, coronary artery disease, diabetes mellitus, hypertension, myocardial infarction and respiratory disease, and whether or not a patient was treated with melatonin, quetiapine, trazodone and benzodiazepines after being intubated. Being at least 65 years old (HR: 1.78, 95% CI: 1.36 - 2.32, p-value = 2.35E-05), having a history of chronic kidney disease (HR: 6.32, 95% CI: 3.54 - 10.3, p-value = 4.32E-10) were significantly associated with a negative outcome in intubation periods for COVID-19 patients (Table 2). Having a history of respiratory disease (HR: 0.493, 95% CI: 0.367 - 0.663, p-value = 2.78E-06) and exposure to quetiapine after intubation (HR: 0.289, 95% CI: 0.210 - 0.398, p-value = 2.37E-14), benzodiazepines after intubation (HR: 0.585, 95% CI: 0.439 - 0.78, p-value = 2.30E-04) and melatonin after intubation (HR: 0.131, 95% CI: 7.76E-02 - 0.223, p-value = 8.19E-14) are significantly associated with a positive outcome in intubation periods for COVID-19 patients (Table 2).

**Table 2.** Melatonin exposure after intubation multivariate model Cox proportional hazards ratios for intubation periods

| Covariates | COVID19 + |  |  | COVID19 - |  |  | 2018 |  |  |
| --- | --- | --- | --- | --- | --- | --- | --- | --- | --- |
|  | N | Hazards Ratio | P-value | N | Hazards Ratio | P-value | N | Hazards Ratio | P-value |
| Age >= 65 | 409 | 1.78<br>(1.36-2.33) | 2.35E-05 * | 1125 | 1.31<br>(0.887-1.95) | 0.173 | 315 | 2.55<br>(1.77-3.67) | 4.84E-07 * |
| Melatonin exposure after intubation | 196 | 0.131<br>(7.66E-02-0.223) | 8.19E-14 * | 319 | 0.278<br>(0.142-0.542) | 1.72E-04 * | 187 | 0.423<br>(0.274-0.653) | 1.03E-04 * |
| Race and Ethnicity |  |  |  |  |  |  |  |  |  |
| American Indian or Alaskan Native | 3 | 1.43<br>(0.346-5.93) | 0.621 |  | ± |  |  | ± |  |
| Disease |  |  |  |  |  |  |  |  |  |
| Chronic kidney disease | 671 | 6.32<br>(3.54-11.3) | 4.32E-10 * | 1303 | 6.00<br>(3.67-9.81) | 8.45E-13 * | 259 | 1.40<br>(0.995-1.96) | 5.38E-02 |
| Chronic obstructive lung disease | 85 | 1.35<br>(0.921-1.97) | 0.125 | 244 | 0.765<br>(0.445-1.31) | 0.332 | 187 | 1.10<br>(0.776-1.55) | 0.603 |
| Coronary artery disease | 145 | 1.33<br>(0.937-1.88) | 0.110 | 539 | 0.774<br>(0.473-1.27) | 0.307 | 265 | 0.935<br>(0.640-1.37) | 0.729 |
| Diabetes mellitus | 408 | 1.07<br>(0.830-1.38) | 0.603 | 722 | 0.964<br>(0.638-1.46) | 0.864 | 283 | 1.25<br>(0.880-1.78) | 0.213 |
| Hypertension | 415 | 0.987<br>(0.754-1.29) | 0.922 | 1181 | 1.04<br>(0.679-1.58) | 0.867 | 471 | 1.00<br>(0.690-1.44) | 0.991 |
| Myocardial infarction | 80 | 0.978<br>(0.645-1.48) | 0.916 | 216 | 3.22<br>(1.95-5.33) | 5.13E-06 * | 160 | 1.33<br>(0.901-1.95) | 0.152 |
| Respiratory Disease | 706 | 0.493<br>(0.367-0.663) | 2.78E-06 * | 1819 | 1.02<br>(0.685-1.51) | 0.936 |  | ± |  |
| Quetiapine exposure after intubation | 252 | 0.289<br>(0.210-0.398) | 2.37E-14 * | 119 | 0.557<br>(0.293-1.06) | 7.54E-02 | 217 | 0.520<br>(0.358-0.756) | 6.06E-04 * |
| Trazodone exposure after intubation | 20 | 0.231<br>(3.20E-02-1.67) | 0.146 | 55 | 0.772<br>(0.241-2.47) | 0.662 | 49 | 0.763<br>(0.379-1.54) | 0.451 |
| Benzodiazepines exposure after intubation | 729 | 0.585<br>(0.439-0.778) | 2.30E-04 * | 2484 | 0.358<br>(0.250-0.513) | 1.95E-08 * | 611 | 1.17<br>(0.762-1.79) | 0.477 |

± Not determined

Having a history of chronic kidney disease (HR: 5.14, 95% CI: 3.18 - 8.29, p-value = 2.08E-11) and myocardial infarction (HR: 3.22, 95% CI: 1.95 - 5.33, p-value = 5.13E-06) were significantly associated with a negative outcome following intubation (Table 2). Exposure to benzodiazepines after intubation (HR: 0.358, 95% CI: 0.250 - 0.513, p-value = 1.95E-08) and exposure to melatonin after intubation (HR: 0.278, 95% CI: 0.142 - 0.542, p-value = 1.72E-04) were significantly associated with a positive outcome for non-COVID-19 patients (Table 2). Being at least 65 years old (HR: 2.55, 95% CI: 1.77 - 3.67, p-value = 4.84E-07) was significantly associated with a negative outcome in intubation periods from 2018 (Table 2). Exposure to quetiapine after intubation (HR: 0.520, 95% CI: 0.358 - 0.756, p-value = 6.06E-04) and melatonin after intubation (HR: 0.423, 95% CI: 0.274 - 0.653, p-value = 1.03E-03) were significantly associated with a positive outcome in intubation periods from 2018 (Table 2).

### Melatonin treatment is associated with increased survival among COVID-19+ patients requiring mechanical ventilation

In a follow up analysis of the intubation periods where the patient required mechanical ventilation, we fit a Cox proportional hazards model using age (binary for whether the patients was older than 65), whether or not the patient self-identified as Black or African American, whether or not the patient had a history of asthma, chronic kidney disease, chronic obstructive lung disease and respiratory disease and whether or not a patients was treated with melatonin, quetiapine, trazodone and benzodiazepines after intubation. Having a history of chronic kidney disease (HR: 3.00, 95% CI: 1.07 - 8.45, p-value = 3.71E-02) was significantly associated with a negative outcome in intubation periods for COVID-19 patients (Table 3). Self identifying as Black or African American (HR: 0.403, 95% CI: 0.193 - 0.839, p-value - 1.15E-02), having a history of respiratory disease (HR: 0.433, 95% CI: 0.250-0.749, p-value = 2.80E-03) and exposure to quetiapine after intubation (HR: 0.404, 95% CI: 0.262 - 0.624, p-value = 4.31E-05), benzodiazepines after intubation (HR: 0.329, 95% CI: 0.187 - 0.580, p-value = 1.19E-04) and melatonin after intubation (HR: 0.127, 95% CI: 6.01E-02 - 0.269, p-value = 7.15E-08) after intubation were significantly associated with a positive outcome for intubation periods for COVID-19 patients where mechanical ventilation was required (Table 3).

**Table 3.** Melatonin exposure after intubation multivariate model Cox proportional hazards ratios for intubation periods requiring mechanical ventilation

| Covariates | COVID19 + |  |  | COVID19 - |  |  | 2018 |  |  |
| --- | --- | --- | --- | --- | --- | --- | --- | --- | --- |
|  | N | Hazards Ratio | P-value | N | Hazards Ratio | P-value | N | Hazards Ratio | P-value |
| Age >= 65 | 144 | 1.51<br>(0.947-2.40) | 8.33E-02 | 90 | 3.07<br>(1.39-6.78) | 5.54E-03 * | 273 | 3.06<br>(2.10-4.45) | 5.44E-09 * |
| Melatonin exposure after intubation | 112 | 0.127<br>(6.01E-02-0.269) | 7.15E-08 * | 34 | 0.689<br>(0.275-1.72) | 0.425 | 152 | 0.492<br>(0.315-0.768) | 1.78E-03 * |
| Race and Ethnicity |  |  |  |  |  |  |  |  |  |
| Black or African American | 62 | 0.403<br>(0.193-0.839) | 1.15E-02 * | 30 | 0.988<br>(0.373-2.61) | 0.980 | 96 | 0.915<br>(0.587-1.43) | 0.696 |
| Disease |  |  |  |  |  |  |  |  |  |
| Asthma | 32 | 0.452<br>(0.138-1.38) | 0.190 | 15 | 1.16<br>(0.255-5.25) | 0.849 | 132 | 0.828<br>(0.526-1.30) | 0.415 |
| Chronic kidney disease | 270 | 3.14<br>(1.12-8.80) | 2.98E-02 * | 127 | 1.55<br>(0.693-3.48) | 0.285 | 223 | 1.44<br>(1.02-2.04) | 3.86E-02 * |
| Chronic obstructive lung disease | 20 | 1.84<br>(0.923-3.66) | 8.32E-02 | 18 | 1.17<br>(0.386-3.53) | 0.785 | 163 | 1.20<br>(0.816-1.75) | 0.360 |
| Respiratory Disease | 275 | 0.433<br>(0.250-0.749) | 2.80E-03 * | 174 | 0.891<br>(0.414-1.92) | 0.768 |  | ± |  |
| Quetiapine exposure after intubation | 175 | 0.404<br>(0.262-0.624) | 4.31E-05 * | 39 | 0.412<br>(0.156-1.09) | 7.42E-02 | 181 | 0.482<br>(0.322-0.722) | 4.04E-04 * |
| Trazodone exposure after intubation | 14 | 0.334<br>(4.48E-02-2.50) | 0.286 | 9 | 0.766<br>(0.163-3.59) | 0.735 | 39 | 0.871<br>(0.414-1.83) | 0.716 |
| Benzodiazepines exposure after intubation | 272 | 0.329<br>(0.187-0.580) | 1.19E-04 * | 163 | 0.754<br>(0.383-1.49) | 0.415 | 519 | 1.28<br>(0.809-2.02) | 0.291 |

± Not determined

Being at least 65 years old (HR: 3.07, 95% CI: 1.39 - 6.78, p-value = 5.54E-03) was significantly associated with a negative outcome in intubation periods for non-COVID-19 patients where mechanical ventilation was required (Table 3). Being at least 65 years old (HR: 3.06, 95% CI: 2.10 - 4.45, p-value = 5.44E-09) and having a history of chronic kidney disease (HR: 1.44, 95% CI: 1.02 - 2.04, p-value = 3.86E-02) were significantly associated with a negative outcome in intubation periods from 2018 (Table 3). Exposure to quetiapine after intubation (HR: 0.482, 95% CI: 0.322 - 0.722, p-value = 4.04E-04) and melatonin after intubation (HR: 0.492, 95% CI: 0.315 - 0.768, p-value = 1.78E-03) were associated with a positive outcome in intubation periods from 2018 (Table 3).

### Chart review of COVID-19 patients treated with melatonin

Following the consistent significant associations between melatonin exposure following intubation and a positive outcome in intubation periods and intubation periods requiring mechanical ventilation for COVID-19 patients, we were interested in the clinical nature of the melatonin prescription. We conducted a manual chart review of 50 randomly identified intubated COVID-19 patients to identify the justification, if any, for melatonin treatment. Of the 34 patients with justifications accompanying melatonin prescription, 21 patients’ charts referenced insomnia, sleep wake cycle or difficulty sleeping for melatonin being prescribed and 18 patients’ charts referenced anxiety, delirium, agitation or agitation delirium (Table 4). Additionally, five patients’ charts referenced sedation, three patients’ charts referenced derangement, altered mental status or mood, and 1 patient’s chart referenced each difficulty waning sedation, adjuvant for presentation of respiratory disorder and pain (Table 4).

**Table 4.** Frequency of terms associated with melatonin treatment

| **Terms** | **N** | **% (of 34)** |
| --- | --- | --- |
| **Derangement, Altered mental status, Mood** | 3 | 8.82 |
| **Insomnia, Sleep wake cycle, Difficulty sleeping** | 21 | 61.8 |
| **Anxiety, Delirium, Agitation, Agitated delirium** | 18 | 52.9 |
| **Difficulty weaning sedation** | 1 | 2.94 |
| **Sedation** | 5 | 14.7 |
| **Adjuvant for presentation of respiratory disorder** | 1 | 2.94 |
| **Pain** | 1 | 2.94 |

## Discussion

In our retrospective analysis of patients who sought care at NYP/CUIMC between February 1st, 2020 and August 1st, 2020, we investigated the effects of hormone exposure in patients requiring endotracheal intubation on mortality.

For the 948 intubation periods among 791 patients who were diagnosed with COVID-19 or infected with SARS-CoV2, there was exposure data for 14 hormone drugs prior to intubation: insulin glargine, insulin human, dronabinol, hydrocortisone, triamcinolone, budesonide, melatonin, dexamethasone, vasopressin, prednisone, methylprednisolone, levothyroxine, fludrocortisone and insulin lispro (Table S1). Additionally, there was exposure data for 17 hormone drugs following intubation: insulin glargine, insulin human, dronabinol, hydrocortisone, triamcinolone, budesonide, melatonin, dexamethasone, vasopressin, prednisone, methylprednisolone, levothyroxine, salmon calcitonin, fludrocortisone, insulin lispro, desmopressin and clobetasol (Table S1).

Univariate survival analysis identified exposure to insulin glargine, budesonide, melatonin, prednisone, methylprednisolone and insulin lispro after intubation as significantly associated with a positive outcome following intubation and exposure to hydrocortisone after intubation is significantly associated with a negative outcome following intubation in intubation periods for COVID-19 patients (Table S3). Additionally, exposure to methylprednisolone and levothyroxine before intubation are significantly associated with a negative outcome following intubation in intubation periods requiring mechanical ventilation for COVID-19 patients (Table S2). Exposure to hydrocortisone, methylprednisolone and levothyroxine after intubation are significantly associated with a negative outcome following intubation in intubation periods requiring mechanical ventilation for COVID-19 patients and exposure to melatonin after intubation is significantly associated with a positive outcome (Table S3).

Univariate survival analysis also identified age (as a continuous variable and binary variable), self-identifying as American Indian or Alaskan Native and a history of chronic kidney disease, chronic obstructive lung disease, coronary artery disease, diabetes mellitus, hypertension and myocardial infarction as significantly associated with a negative outcome following intubation in intubation periods for COVID-19 patients (Table S2). A history of respiratory disease was significantly associated with a positive outcome following intubation (Table S2). Age (as a continuous variable and binary variable) and a history of chronic kidney disease and chronic obstructive lung disease are significantly associated with a negative outcome following intubation in intubation periods requiring mechanical ventilation for COVID-19 patients (Table S2). Having a history of asthma and respiratory disease are significantly associated with a positive outcome following intubation (Table S2).

Treatment with dexamethasone following intubation was associated, though not significantly, with a positive outcome in our univariate analysis of intubation periods for COVID-19 patients (Table S3). Furthermore, the association, though not significant, was observed in intubation periods for COVID-19 and non-COVID-19 patients when accounting for other covariates (Table S4). However, our analysis did not indicate an association between dexamethasone treatment following intubation and positive outcome in intubation periods requiring mechanical ventilation unlike the observation from the RECOVERY Collaborative Group’s study[15] (Table S5). The power of our analysis is most likely limited due to the small sample size (N=4)

Moreover, our results identify exposure to melatonin as significantly associated with a positive outcome after intubation in univariate analyses of intubation periods for COVID-19 patients and intubation periods where mechanical ventilation was required for COVID-19 patients (Table S3) concurring with previous studies on the attenuation of cardiovascular responses following anesthesia[17], duration of mechanical ventilation in hemorrhagic stroke patients[18], and identification of melatonin acting as a regulator of inflammation[19]. The significant association of exposure to melatonin following intubation with a positive outcome in a multivariate model of COVID-19 and non-COVID-19 patients suggests that melatonin exposure is not specifically attenuating inflammation due to SARS-CoV2 infection (Table 2). However, the multivariate model focusing on intubation periods where mechanical ventilation was required indicated that exposure to melatonin was only only associated with a positive outcome in COVID-19 patients suggesting that melatonin’s mechanism of action in the most severe cases of COVID-19 may be targeted to SARS-CoV2 induced inflammation (Table 3).

A manual chart review of a subset of intubated COVID-19 patients did not reveal any inflammation specific goals for the treatment (Table 4). While melatonin is a popular over-the-counter sleep aid, our results lend support to the need for further follow-up into the mechanism of action of how melatonin may attenuate inflammation and specifically more studies into the observed association in severely affected COVID-19 patients.

## Methods

### Ethics statement

The study is approved by the Columbia University Irving Medical Center Institutional Review Board (IRB# AAAL0601) and the requirement for an informed consent was waived. A data request associated with this protocol was submitted to the Tri-Institutional Request Assessment Committee (TRAC) of New-York Presbyterian, Columbia, and Cornell and approved.

### Cohort identification

We collected the data electronic health records for 189,987 patients who sought care at NYP/ CUIMC between February 1st, 2020 and August 1st, 2020. From those patients, we identified a cohort of 13,394 patients who tested positive for SARS-CoV2 infection using a nasopharyngeal RT-PCR test or were clinically diagnosed with COVID-19. From those patients we identified 948 intubation periods for 791 patients who required endotracheal intubation. We identified 3,497 intubation periods for 2,981 patients who required endotracheal intubation but were not diagnosed with COVID-19. We also identified 747 intubation periods for 637 patients, who sought care at NYP/CUIMC between February 1st, 2018 and August 1st, 2018, requiring endotracheal intubation from our CDW.

### Identify intubation-extubation periods and ventilator use

To begin, we used hospital admission and discharge data to identify visits lasting more than one day to remove any patients who were admitted for outpatient procedures. We identified intubation orders using the display name and description of orders that had been completed - we used a similar method to identify extubation orders - and filtered for visits where the intubation date and discharge date were not the same to remove any patients who were intubated during a surgical procedure. We used the dates of extubation orders to define the end of each intubation-extubation period for each patient and identified the start of each intubation-extubation period as the first intubation order following any previous extubation order. Intubation-extubation periods beginning before February 1st, 2020 (i.e. those with an extubation order, but missing an intubation order), were excluded from the analysis. Intubations that are not followed by an extubation were censored up to the discharge date or the final date of analysis, August 1st, 2020. For patients who died within seven days following extubation, intubation-extubation periods were censored up to the death date.

For the 2018 intubation periods, patients, who had been intubated, were identified using procedures identified as ‘Intubation, endotracheal, emergency procedure’, ‘Insertion of Endotracheal Airway into Trachea, Via Natural or Artificial Opening’, or ‘Insertion of Endotracheal Airway into Trachea, Via Natural or Artificial Opening Endoscopic’. We identified days on which patients were intubated using procedures identified as ‘’Unlisted procedure, larynx’ or ‘Subsequent hospital care, per day, for the evaluation and management’. We used the dates of intubation procedures to define the start of each intubation-extubation period and the maximum date of intubation care following the intubation procedure as the end of each intubation-extubation period.

For the 2020 intubation periods, patients requiring the use of a ventilator were identified using the display name and description of order that had not been cancelled such that the order date occurred during the intubation-extubation period. Intubation-extubation periods for each patient with mechanical ventilation order were coded as 1, while those without were coded as 0. For the 2018 intubation periods, patients requiring the use of a ventilator were identified as “Respiratory Ventilation, Less than 24 Consecutive Hours”, “Respiratory Ventilation, 24-96 Consecutive Hours”, and “Respiratory Ventilation, Greater than 96 Consecutive Hours”. Intubation-extubation periods for each patient with a ventilation order were coded as 1, while those without were coded as 0.

### Identify demographic information

For each patient within our cohorts of interest, we identified the patient’s reported birth date, death date, if the patient had died, sex, race(s) and ethnicity. We calculated the age of each patient on their first admission to the hospital within the observation period. For sex, patients identifying their sex as male were identifed as 1, while those who reported their sex as female or other were coded as 0. For ethnicity, patients who reported their ethnicity as Hispanic or Latino or Spanish origin were coded as 1, while those who reported their ethnicity as Not Hispanic or Latino or Spanish origin or who did not report their ethnicity were coded as 0. For each possible racial group: American Indian or Alaskan Native, Native Hawaiian or Other Pacific Islander, Ashkenazi Jewish, Black or African American, Asian, and White, patients were coded as 1 if they reported identifying as a member of that racial group.

Among intubation periods for COVID-19 patients, increasing age, as a continuous variable (HR: 1.05, 95% CI: 1.04 −1.06, p-value = 4.42E-24) and as a binary variable of age greater than or equal to 65 years (HR: 3.25, 95% CI: 2.52 - 4.9, p-value = 1.33E-19), and self-identifying as (HR: 4.36, 95% CI: 1.08 - 17.6, p-value = 3.80E-02) was significantly associated with a negative outcome (Table S1 and Figure S1). Among the subset of intubation periods where mechanical ventilation was required for COVID-19 patients increasing age, as a continuous variable (HR: 1.04, 95% CI: 1.02 −1.06, p-value = 1.39E-5) and as a binary variable of age greater than or equal to 65 (HR: 2.85, 95% CI: 1.84 - 4.40, p-value = 2.42E-06) was significantly associated with a negative outcome(Table S1 and Figure S3). Conversely, self-identifying race as Black or African American (HR: 0.347, 95% CI: 0.175 - 0.689, p-value = 2.49E-3) was significantly associated with a positive outcome (Table S1 and Figure S3).

### Identifying patient comorbidities

For patients in each cohort, we used the data available in the EHR at NYP/CUIMC and in the CDW at NYP/CUIMC to identify whether of not a patient had a history of asthma, cardiovascular disease, chronic kidney disease, chronic obstructive lung disease, coronary artery disease, delirium, diabetes mellitus, diabetic nephropathy, diabetic neuropathy, diabetic retinopathy, diabetic vasculopathy, heart failure, hypertension, insomnia, myocardial infarction, obesity, and respiratory disorder using ICD-10 diagnosis codes in the EHR and SNOMED-CT codes and relationships in data from the CDW. For each disease in our survival analysis, patients with a history were coded as 1, while those without a history were coded as 0.

Among intubation periods for COVID-19, having a history of chronic kidney disease (HR: 5.90, 95% CI: 3.40 - 10.4, p-value = 3.73E-10), chronic obstructive lung disease (HR: 1.82, 95% CI: 1.29 - 2.58, p-value = 7.37E-4), coronary artery disease (HR: 1.65, 95% CI: 1.23 - 2.21, p-value = 8.41E-04), diabetes mellitus (HR: 1.61, 95% CI: 1.27 - 2.04, p-value =8.83E-05), hypertension (HR: 1.62, 95% CI: 1.20 - 1.93, p-value = 5.20E-04) and myocardial infarction (HR: 1.56, 95% CI: 1.07 - 2.27, p-value = 1.96E-02) were associated with a negative outcome. Having a history of respiratory disease (HR: 0.630, 95% CI: 0.480 - 0.830, p-value = 1.06E-30) was significantly associated with a positive outcome following intubation (Table S1 and Figure S1).

Among the subset of intubation periods where mechanical ventilation was required for COVID-19, having a history of chronic kidney disease (HR: 3.63, 95% CI: 1.32 - 9.89, p-value = 1.17E-02) and chronic obstructive lung disease (HR: 2.06, 95% CI: 1.07 - 3.98, p-value = 3.11E-02) were significantly associated with a negative outcome (Table S2 and Figure S3). Conversely, having a history of asthma (HR: 0.299, 95% CI: 9.50E-02 - 0.950, p-value = 4.00E-2) and respiratory disease (HR: 0.457, 95% CI: 0.273 - 0.766, p-value = 2.98E-3) were significantly associated with a positive outcome (Table S1 and Figure S3).

### Identify patient drug treatments

For the 2020 intubation periods, we identified the drug names, the associated Nation Library of Medication RXNorm identification code and the time of order from orders that had been completed or time of action from medication administration record. We then mapped RXNorm codes to DrugBank codes and utilized the associated DrugBank categories to identified drugs classified as hormones. Patients were considered as being treated with a drug before intubation if they had at least one completed order or administration between February 1st, 2020 and the start of an intubation-extubation period outside of any other intubation-extubation period. Similarly, patients were considered as being treated with a drug after intubation if they had at least one completed order or administration on or after the start of the intubation-extubation period through the end of the period.

For the intubation periods from 2018, we identified drug names, the associated RX Norm identification code, and the start date and end date of their drug regiment censored between February 1st, 2018 and August 1st, 2018. Patients were considered as being treated with a drug before intubation if any part of the treatment period occurred between the visit start day and the the intubation day outside of any other intubation-extubation period. Similarly patients were were considered as being treated with a drug after intubation if any part of the treatment period occurred between the intubation day and the extubation day or the censoring date.

### Identifying patient outcomes

For patients with a single intubation-extubation period, we identified patients for whom intubation was not beneficial as those who died within the intubation-extubation period or within seven days following the end of that period. For patients with multiple intubation-extubation periods, we identified patients for whom intubation was not beneficial as those who died within the final intubation-extubation period or within seven days following the end of that period. For our survival analysis, intubation-extubation periods where the patient did not die within seven days of extubation were coded as 0, while those who died within seven days were coded as 1. For intubation-extubation periods that did not result in death within seven days, time to event was equal to the length of the intubation-extubation period. For intubation-extubation periods that did not result in death within seven days, time to event was equal to the length of time from the intubation day to the death of the patient.

### Statistical modeling and software

We used Jupyter Notebooks (jupyter-client version 5.3.4 and jupyter-core version 4.6.1) running Python 3.7 and all fit models using the python lifelines package (version 0.24.4). We used MySQL and python libraries (pymysql, numpy, pandas and pickle) to extract and prepare the data for modeling.

## Data Availability

All scripts used for data preparation and analysis are available from GitHub as a Jupyter Notebook (https://github.com/tatonetti-lab/melatonin).

## Acknowledgements

This work was funded by US National Institutes of Health grant R35GM131905 to NT.

---

**Figure S1.**
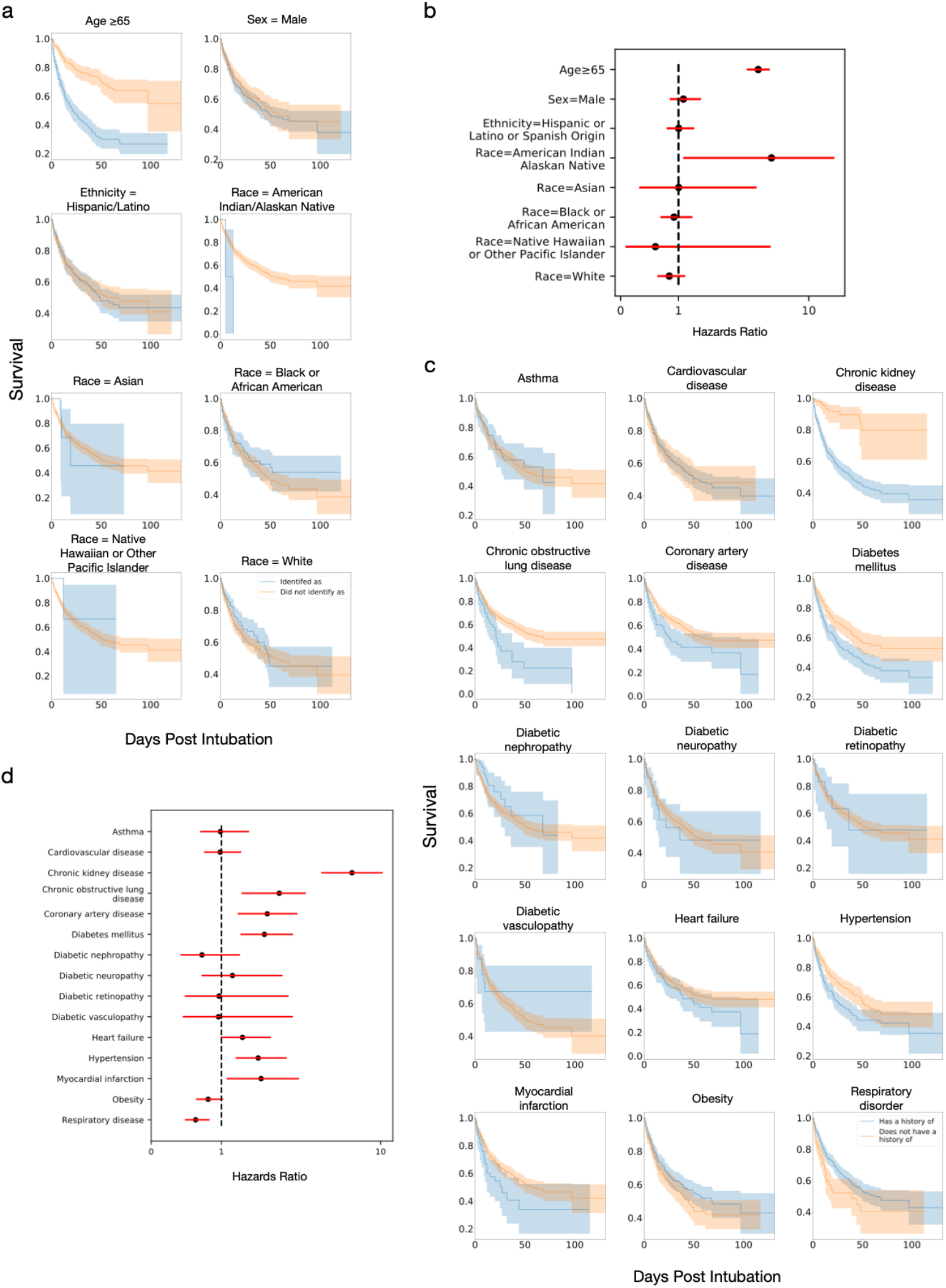
Kaplan-Meier curves for (a) demographic and (c) disease covariates for COVID-19 intubation periods. Comparison of hazard rations for (b) demographic and (d) disease covariates shown in (a) and (c), respectively.

**Figure S2.**
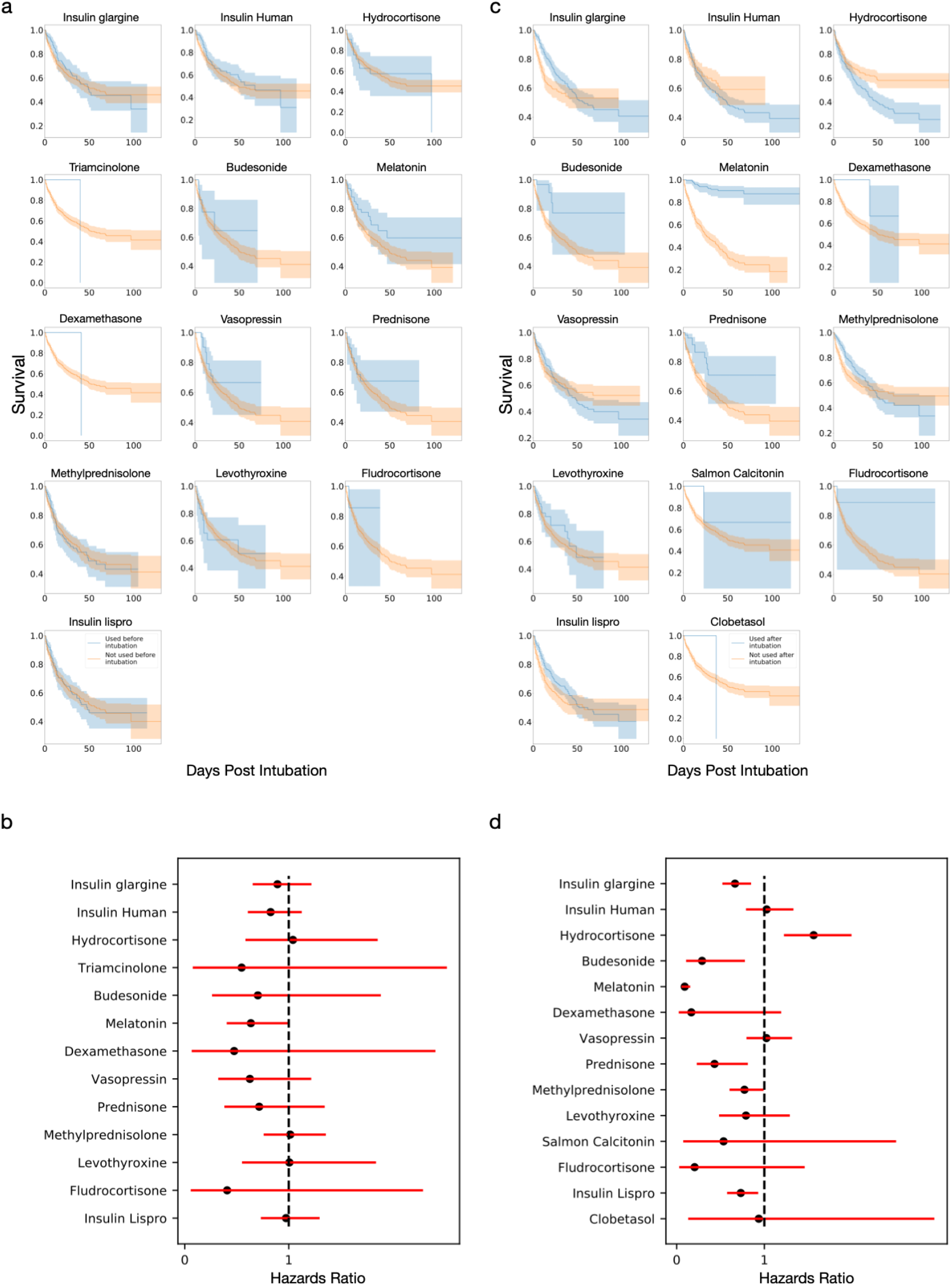
Kaplan-Meier curves for hormones exposure (a) before and (c) after intubation for COVID-19 intubation periods. Comparison of hazard rations for (b) demographic and (d) disease covariates shown in (a) and (c), respectively.

**Figure S3.**
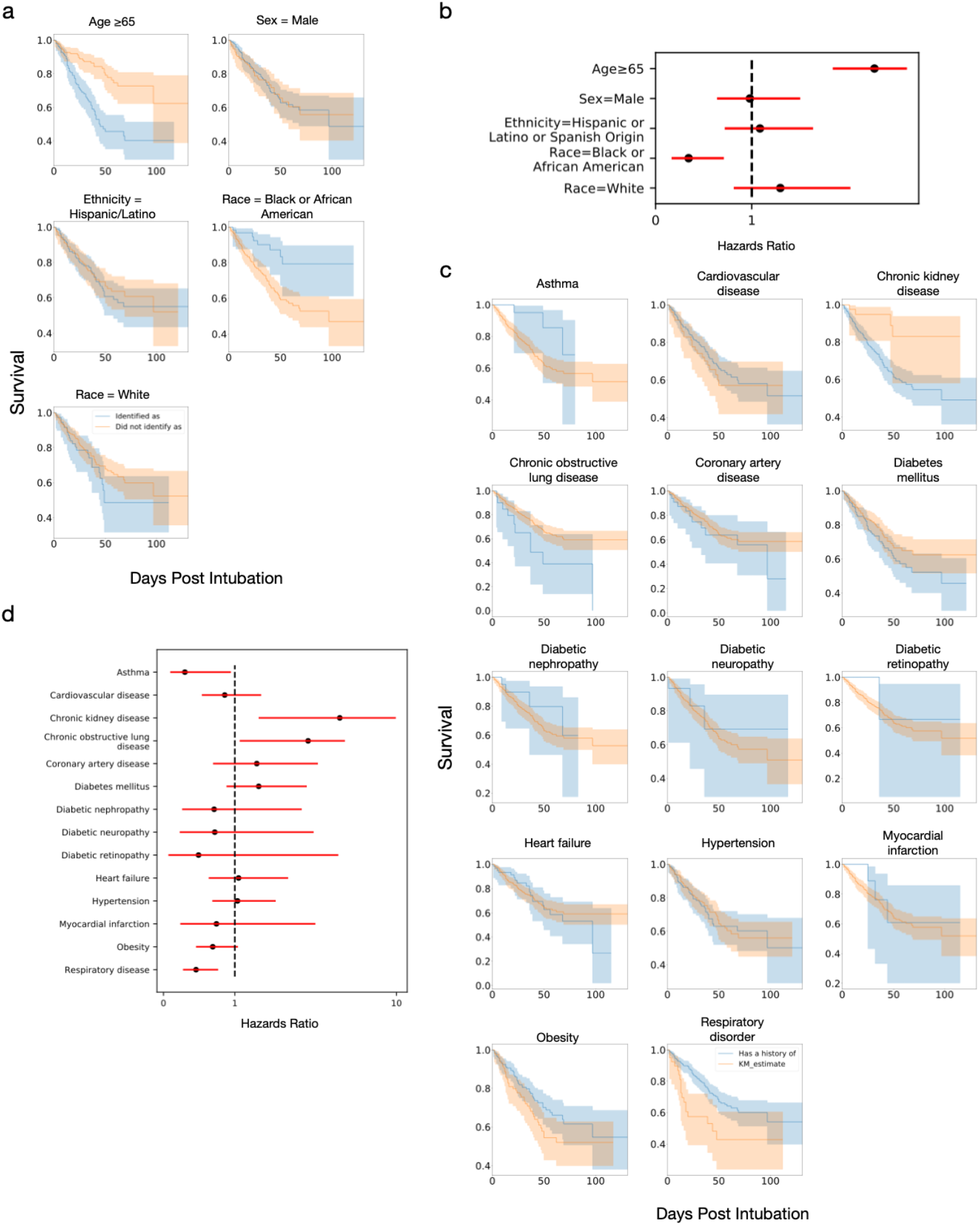
Kaplan-Meier curves for (a) demographic and (c) disease covariates for COVID-19 intubation periods requiring mechanical ventilation. Comparison of hazard rations for (b) demographic and (d) disease covariates shown in (a) and (c), respectively.

**Figure S4.**
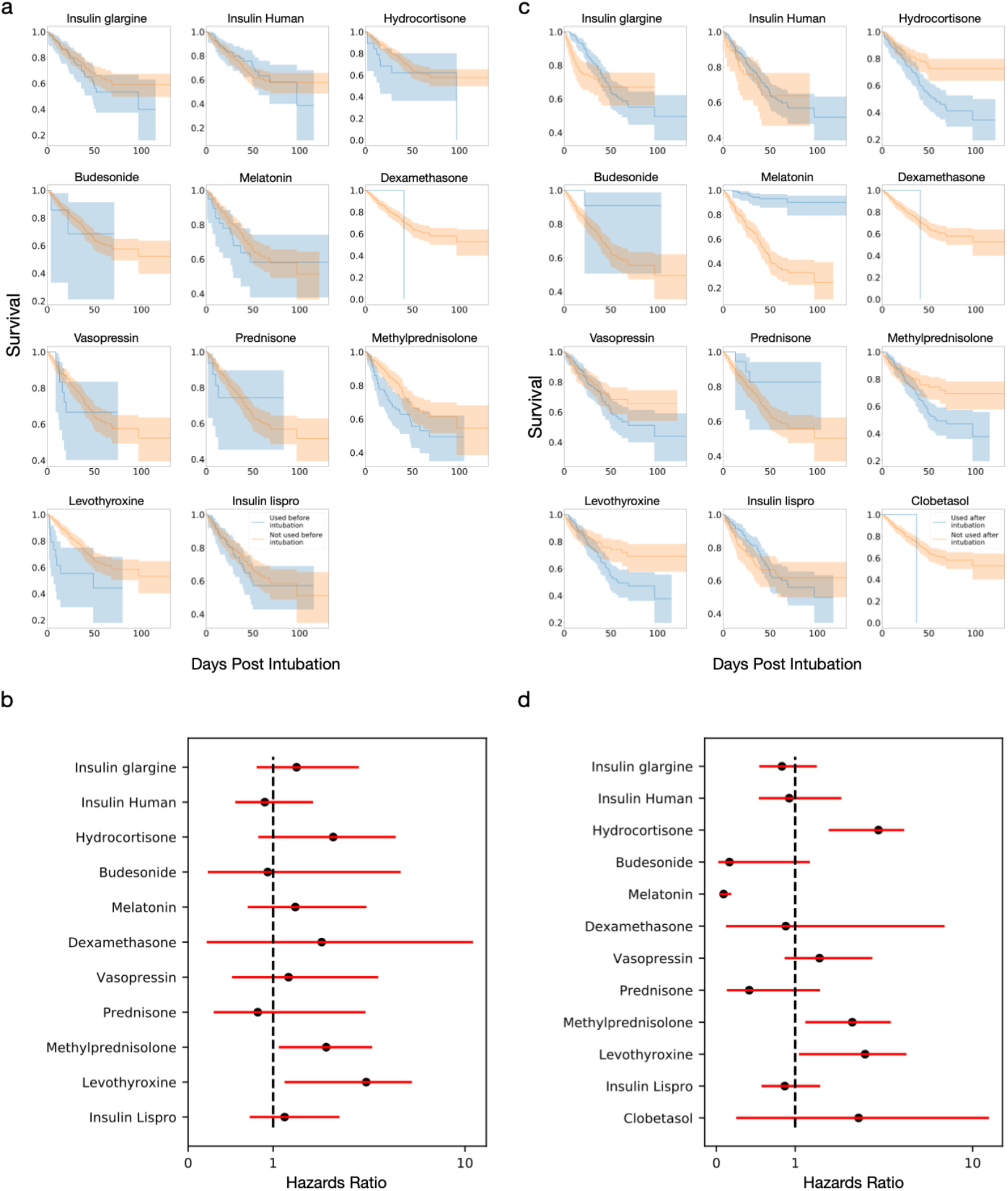
Kaplan-Meier curves for hormones exposure (a) before and (c) after intubation for COVID-19 intubation periods requiring mechanical ventilation. Comparison of hazard rations for (b) demographic and (d) disease covariates shown in (a) and (c), respectively.

**Table S1.** Frequency of drug exposure in intubation periods

|  | COVID19 + | COVID19 - | 2018 |
| --- | --- | --- | --- |
| **Quetiapine before intubation** | 52<br>5.49% | 53<br>1.52% | 77<br>10.31% |
| **Quetiapine after intubation** | 252<br>26.58% | 119<br>3.40% | 217<br>29.05% |
| **Trazodone before intubation** | 13<br>1.37% | 28<br>0.80% | 27<br>3.61% |
| **Trazodone after intubation** | 20<br>2.11% | 55<br>1.57% | 49<br>6.56% |
| **Benzodiazepines before intubation** | 174<br>18.35% | 544<br>15.56% | 518<br>69.34% |
| **Benzodiazepines after intubation** | 729<br>76.90% | 2484<br>71.03% | 611<br>81.79% |
| **Insulin glargine before intubation** | 138<br>14.56% | 129<br>3.69% | ± |
| **Insulin glargine after intubation** | 341<br>35.97% | 273<br>7.81% | ± |
| **Insulin Human before intubation** | 150<br>15.82% | 162<br>4.63% | ± |
| **Insulin Human after intubation** | 486<br>51.27% | 432<br>12.35% | ± |
| **Dronabinol before intubation** | ≤10<br>≤1.05% | ≤10<br>≤0.27% | ± |
| **Dronabinol after intubation** | ≤10<br>≤1.05% | 12<br>0.34% | ± |
| **Hydrocortisone before intubation** | 34<br>3.59% | 74<br>2.12% | ± |
| **Hydrocortisone after intubation** | 196<br>20.68% | 198<br>5.66% | ± |
| **Triamcinolone before intubation** | ≤10<br>≤1.05% | ≤10<br>≤0.27% | ± |
| **Triamcinolone after intubation** | 15<br>1.58% | 23<br>0.66% | ± |
| **Budesonide before intubation** | 18<br>1.90% | 67<br>1.92% | ± |
| **Budesonide after intubation** | 33<br>3.48% | 108<br>3.09% | ± |
| **Melatonin before intubation** | 86<br>9.07% | 172<br>4.92% | 93<br>12.45% |

Table S1 Frequency of drug exposure in intubation periods, cont.
|  | COVID19 + | COVID19 - | 2018 |
| --- | --- | --- | --- |
| **Melatonin after intubation** | 196<br>20.68% | 319<br>9.12% | 187<br>25.03% |
| **Dexamethasone before intubation** | ≤10<br>≤1.05% | 67<br>1.92% | 78<br>10.44% |
| **Dexamethasone after intubation** | 18<br>1.90% | 206<br>5.89% | 81<br>10.84% |
| **Vasopressin before intubation** | 37<br>3.90% | 73<br>2.09% | ± |
| **Vasopressin after intubation** | 221<br>23.31% | 283<br>8.09% | ± |
| **Prednisone before intubation** | 41<br>4.32% | 98<br>2.80% | ± |
| **Prednisone after intubation** | 60<br>6.33% | 171<br>4.89% | ± |
| **Methylprednisolone before intubation** | 146<br>15.40% | 104<br>2.97% | ± |
| **Methylprednisolone after intubation** | 286<br>30.17% | 315<br>9.01% | ± |
| **Levothyroxine before intubation** | 36<br>3.80% | 78<br>2.23% | ± |
| **Levothyroxine after intubation** | 63<br>6.65% | 201<br>5.75% | ± |
| **Salmon Calcitonin after intubation** | ≤10<br>≤1.05% | ≤10<br>≤0.27% | ± |
| **Fludrocortisone before intubation** | ≤10<br>≤1.05% | ≤10<br>≤0.27% | ± |
| **Fludrocortisone after intubation** | ≤10<br>≤1.05% | 15<br>0.43% | ± |
| **Insulin Lispro before intubation** | 172<br>18.14% | 237<br>6.78% | ± |
| **Insulin Lispro after intubation** | 362<br>38.19% | 493<br>14.10% | ± |
| **Desmopressin after intubation** | ≤10<br>≤1.05% | 19<br>0.54% | ± |
| **Clobetasol after intubation** | ≤10<br>≤1.05% | ≤10<br>≤0.27% | ± |

± Not determined

**Table S2.** Demographic and disease univariate Cox proportional hazards ratios for COVID-19 + intubation periods

| Covariates | Intubated Patients |  |  | Mechanically Ventilated Patients |  |  |
| --- | --- | --- | --- | --- | --- | --- |
|  | N | Hazards Ratio | P-value | N | Hazards Ratio | P-value |
| **Age (continous)** | 948 | 1.05<br>(1.04-1.06) | 4.42E-24 * | 315 | 1.04<br>(1.02-1.06) | 1.39E-05 * |
| **Age >= 65** | 409 | 3.25<br>(2.52-4.19) | 1.33E-19 * | 144 | 2.85<br>(1.84-4.40) | 2.42E-06 * |
| **Male** | 577 | 1.09<br>(0.848-1.39) | 0.513 | 196 | 0.980<br>(0.639-1.50) | 0.927 |
| **Race and Ethnicity** |  |  |  |  |  |  |
| **Hispanic or Latin or Spanish origin** | 438 | 1.00<br>(0.793-1.27) | 0.969 | 166 | 1.09<br>(0.719-1.64) | 0.697 |
| **American Indian or Alaskan Native** | 3 | 4.36<br>(1.08-17.6) | 3.80E-02 * |  | ± |  |
| **Asian** | 13 | 1.00<br>(0.321-3.13) | 0.995 |  | ± |  |
| **Black or African American** | 195 | 0.923<br>(0.687-1.24) | 0.595 | 62 | 0.344<br>(0.166-0.710) | 3.92E-03 * |
| **Native Hawaiian or Other Pacific Islander** | 3 | 0.602<br>(8.44E-02-4.29) | 0.612 |  | ± |  |
| **White** | 269 | 0.842<br>(0.634-1.12) | 0.232 | 77 | 1.30<br>(0.813-2.07) | 0.275 |
| **Disease** |  |  |  |  |  |  |
| **Asthma** | 135 | 0.984<br>(0.696-1.39) | 0.929 | 32 | 0.299<br>(9.46E-02-0.945) | 3.98E-02 * |
| **Cardiovascular disease** | 662 | 0.981<br>(0.752-1.28) | 0.891 | 240 | 0.858<br>(0.538-1.37) | 0.521 |
| **Chronic Kidney disease** | 671 | 5.94<br>(3.40-10.4) | 3.73E-10 * | 270 | 3.63<br>(1.33-9.89) | 1.17E-02 * |
| **Chronic obstructive lung disease** | 85 | 1.82<br>(1.29-2.58) | 7.37E-04 * | 20 | 2.06<br>(1.07-3.98) | 3.11E-02 * |
| **Coronary artery disease** | 145 | 1.65<br>(1.23-2.21) | 8.41E-04 * | 32 | 1.31<br>(0.695-2.46) | 0.407 |
| **Diabetes mellitus** | 408 | 1.61<br>(1.27-2.04) | 8.83E-05 * | 138 | 1.33<br>(0.884-2.01) | 0.171 |
| **Diabetic nephropathy** | 56 | 0.723<br>(0.414-1.26) | 0.254 | 21 | 0.710<br>(0.260-1.94) | 0.503 |
| **Diabetic neuropathy** | 52 | 1.16<br>(0.716-1.87) | 0.552 | 15 | 0.719<br>(0.227-2.28) | 0.574 |
| **Diabetic retinopathy** | 28 | 0.966<br>(0.478-1.95) | 0.923 | 7 | 0.492<br>(6.85E-02-3.54) | 0.481 |

Table S2 Demographic and disease univariate Cox proportional hazards ratios for COVID-19 + intubation periods, cont.
| Covariates | Intubated Patients |  |  | Mechanically Ventilated Patients |  |  |
| --- | --- | --- | --- | --- | --- | --- |
|  | N | Hazards Ratio | P-value | N | Hazards Ratio | P-value |
| **Disease** |  |  |  |  |  |  |
| **Diabetic vasculopathy** | 26 | 0.958<br>(0.451-2.03) | 0.910 |  | ± |  |
| **Heart failure** | 198 | 1.30<br>(0.989-1.70) | 5.98E-02 | 60 | 1.05<br>(0.635-1.74) | 0.843 |
| **Hypertension** | 415 | 1.52<br>(1.20-1.93) | 5.20E-04 * | 138 | 1.04<br>(0.684-1.57) | 0.864 |
| **Myocardial infarction** | 80 | 1.56<br>(1.07-2.27) | 1.96E-02 * | 13 | 0.742<br>(0.235-2.35) | 0.612 |
| **Obesity** | 450 | 0.810<br>(0.639-1.03) | 8.10E-02 | 193 | 0.691<br>(0.457-1.04) | 7.87E-02 |
| **Respiratory disease** | 706 | 0.632<br>(0.480-0.832) | 1.06E-03 * | 275 | 0.457<br>(0.273-0.766) | 2.98E-03 * |

± Not determined

**Table S3.**
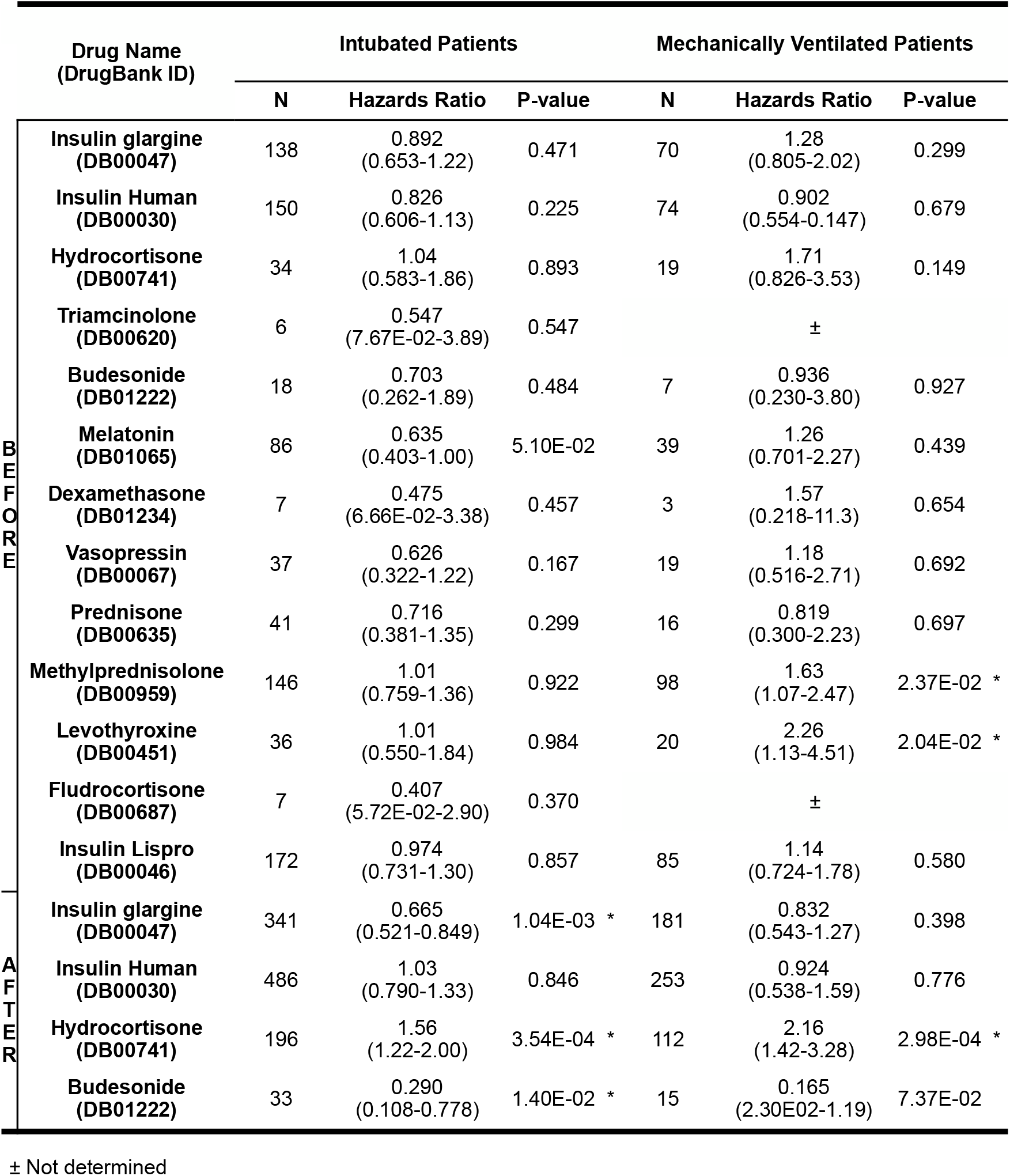
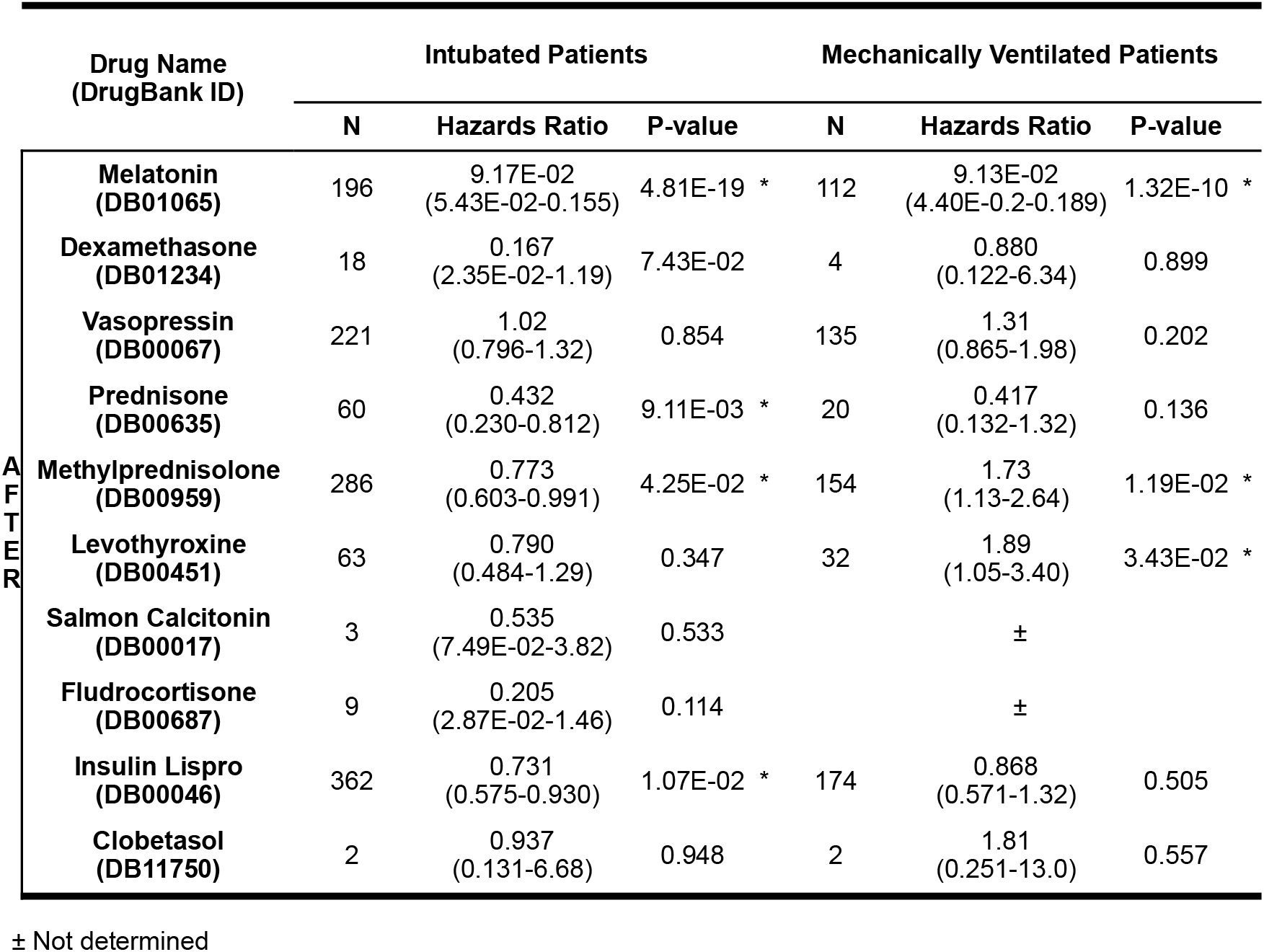
Hormone exposure before and after intubation univariate Cox proportional hazards ratios for COVID-19+ intubation periods

**Table S4.**
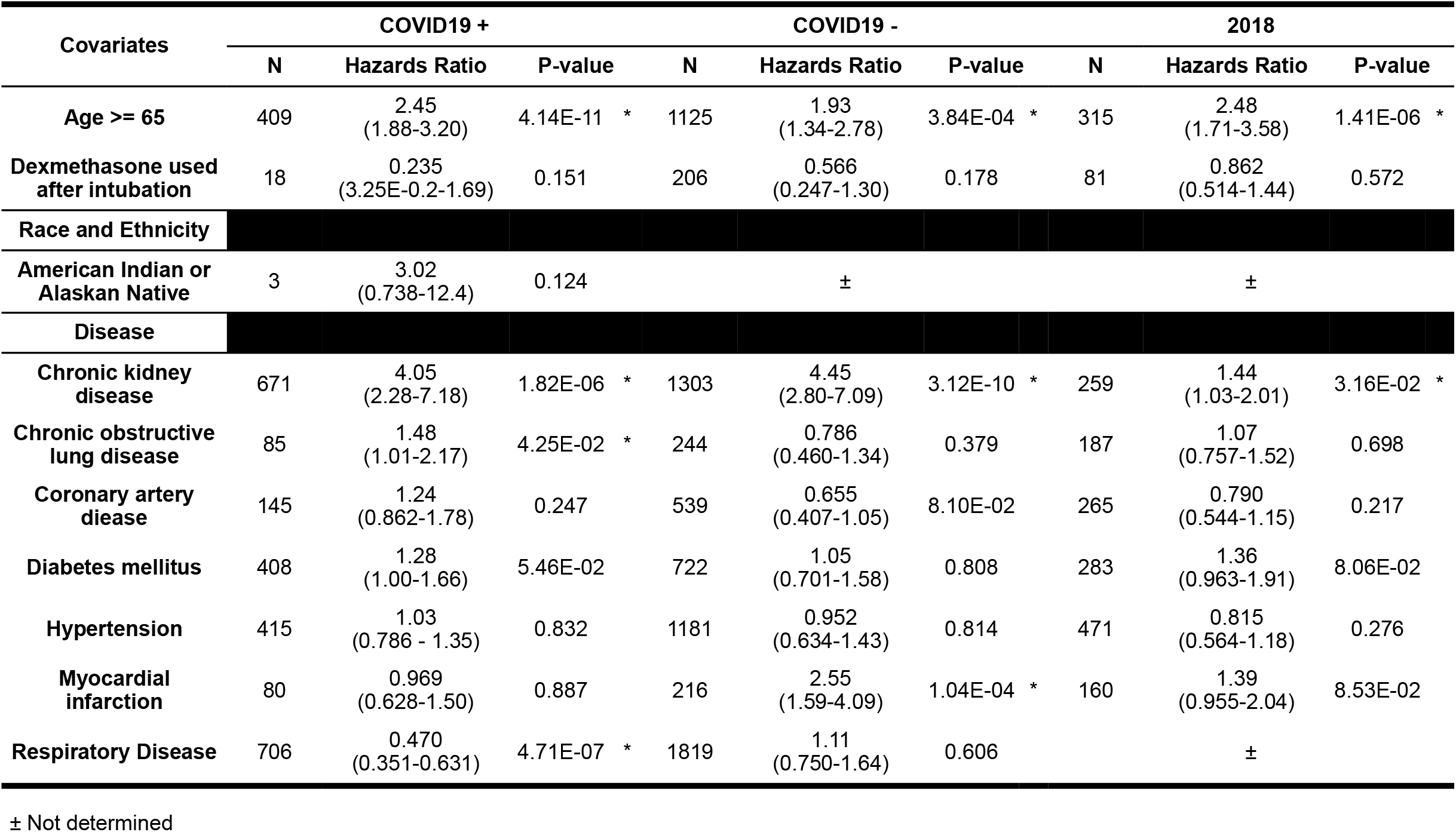
Dexamethasone exposure after intubation multivariate model Cox proportional hazards ratios for intubation periods

| Covariates | COVID19 + |  |  | COVID19 - |  |  | 2018 |  |  |
| --- | --- | --- | --- | --- | --- | --- | --- | --- | --- |
|  | N | Hazards Ratio | P-value | N | Hazards Ratio | P-value | N | Hazards Ratio | P-value |
| **Age >= 65** | 409 | 2.45<br>(1.88-3.20) | 4.14E-11 * | 1125 | 1.93<br>(1.34-2.78) | 3.84E-04 * | 315 | 2.48<br>(1.71-3.58) | 1.41E-06 * |
| **Dexamethasone used after intubation** | 18 | 0.235<br>(3.25E-0.2-1.69) | 0.151 | 206 | 0.566<br>(0.247-1.30) | 0.178 | 81 | 0.862<br>(0.514-1.44) | 0.572 |
| **Race and Ethnicity** |  |  |  |  |  |  |  |  |  |
| **American Indian or Alaskan Native** | 3 | 3.02<br>(0.738-12.4) | 0.124 |  | ± |  |  | ± |  |
| **Disease** |  |  |  |  |  |  |  |  |  |
| **Chronic kidney disease** | 671 | 4.05<br>(2.28-7.18) | 1.82E-06 * | 1303 | 4.45<br>(2.80-7.09) | 3.12E-10 * | 259 | 1.44<br>(1.03-2.01) | 3.16E-02 * |
| **Chronic obstructive lung disease** | 85 | 1.48<br>(1.01-2.17) | 4.25E-02 * | 244 | 0.786<br>(0.460-1.34) | 0.379 | 187 | 1.07<br>(0.757-1.52) | 0.698 |
| **Coronary artery disease** | 145 | 1.24<br>(0.862-1.78) | 0.247 | 539 | 0.655<br>(0.407-1.05) | 8.10E-02 | 265 | 0.790<br>(0.544-1.15) | 0.217 |
| **Diabetes mellitus** | 408 | 1.28<br>(1.00-1.66) | 5.46E-02 | 722 | 1.05<br>(0.701-1.58) | 0.808 | 283 | 1.36<br>(0.963-1.91) | 8.06E-02 |
| **Hypertension** | 415 | 1.03<br>(0.786 - 1.35) | 0.832 | 1181 | 0.952<br>(0.634-1.43) | 0.814 | 471 | 0.815<br>(0.564-1.18) | 0.276 |
| **Myocardial infarction** | 80 | 0.969<br>(0.628-1.50) | 0.887 | 216 | 2.55<br>(1.59-4.09) | 1.04E-04 * | 160 | 1.39<br>(0.955-2.04) | 8.53E-02 |
| **Respiratory Disease** | 706 | 0.470<br>(0.351-0.631) | 4.71E-07 * | 1819 | 1.11<br>(0.750-1.64) | 0.606 |  | ± |  |

± Not determined

**Table S5.** Dexamethasone exposure after intubation multivariate model Cox proportional hazards ratios for intubation periods requiring mechanical ventilation

| Covariates | COVID19 + |  |  | COVID19 - |  |  | 2018 |  |  |
| --- | --- | --- | --- | --- | --- | --- | --- | --- | --- |
|  | N | Hazards Ratio | P-value | N | Hazards Ratio | P-value | N | Hazards Ratio | P-value |
| **Age >= 65** | 144 | 2.29<br>(1.46-3.59) | 3.00E-04 * | 90 | 3.11<br>(1.47-6.55) | 2.91E-03 * | 273 | 2.77<br>(1.90-4.03) | 1.11E-07 * |
| **Dexamethasone used after intubation** | 4 | 0.883<br>(0.121-6.44) | 0.903 | 10 | 0.525<br>(0.124-2.23) | 0.383 | 67 | 0.857<br>(0.494-1.49) | 0.585 |
| **Race and Ethnicity** |  |  |  |  |  |  |  |  |  |
| **Black or African American** | 62 | 0.384<br>(0.185-0.797) | 1.02E-02 * | 30 | 1.03<br>(0.393-2.68) | 0.956 | 96 | 1.00<br>(0.639-1.55) | 0.985 |
| **Disease** |  |  |  |  |  |  |  |  |  |
| **Asthma** | 32 | 0.347<br>(0.109-1.11) | 7.36E-02 | 15 | 1.20<br>(0.267-5.42) | 0.809 | 132 | 0.886<br>(0.566-1.39) | 0.594 |
| **Chronic kidney disease** | 270 | 2.71<br>(0.977-7.52) | 5.56E-02 | 127 | 1.28<br>(0.598-2.73) | 0.526 | 223 | 1.40<br>(0.994-1.98) | 5.39E-02 |
| **Chronic obstructive lung disease** | 20 | 1.96<br>(0.994-3.85) | 5.22E-02 | 18 | 0.923<br>(0.314-2.72) | 0.885 | 163 | 1.06<br>(0.731-1.54) | 0.752 |
| **Respiratory Disease** | 275 | 0.534<br>(0.314-0.910) | 2.11E-02 * | 174 | 0.754<br>(0.358-1.59) | 0.457 |  | ± |  |

± Not determined

**Table S6.** Insomnia and agitation medications univariate Cox proportional hazards ratios for intubation periods

|  | Drug | COVID19 + |  |  | COVID19 - |  |  | 2018 |  |  |
| --- | --- | --- | --- | --- | --- | --- | --- | --- | --- | --- |
|  |  | N | Hazards Ratio | P-value | N | Hazards Ratio | P-value | N | Hazards Ratio | P-value |
| B<br>E<br>F<br>O<br>R<br>E | Melatonin | 86 | 0.635<br>(0.403-1.00) | 5.10E-02 | 172 | 2.20<br>(1.41-3.42) | 4.83E-04 * | 93 | 1.25<br>(0.841-1.85) | 0.273 |
|  | Quetiapine | 52 | 0.536<br>(0.293-0.980) | 4.28E-02 * | 53 | 1.50<br>(0.699-3.22) | 0.298 | 77 | 1.12<br>(0.713-1.75) | 0.632 |
|  | Trazodone | 13 | 0.771<br>(0.287-2.07) | 0.606 | 28 | 1.79<br>(0.570-5.64) | 0.318 | 27 | 0.790<br>(0.349-1.79) | 0.571 |
|  | Benzodiazepines | 174 | 0.477<br>(0.330-0.690) | 8.31E-05 * | 544 | 1.27<br>(0.888-1.82) | 0.190 | 518 | 1.20<br>(0.853-1.68) | 0.300 |
| A<br>F<br>T<br>E<br>R | Melatonin | 196 | 9.17E-02<br>(5.43E-02-0.155) | 4.81E-19 * | 319 | 0.322<br>(0.169-0.615) | 5.94E-04 * | 187 | 0.407<br>(0.269-0.614) | 1.81E-05 * |
|  | Quetiapine | 252 | 0.242<br>(0.178-0.329) | 1.60E-19 * | 119 | 0.838<br>(0.450-1.56) | 0.577 | 217 | 0.471<br>(0.328-0.676) | 4.46E-05 * |
|  | Trazodone | 20 | 8.31E-02<br>(1.17E-02-0.593) | 1.13E-02 * | 55 | 0.697<br>(0.222-2.19) | 0.537 | 49 | 0.527<br>(0.268-1.03) | 6.22E-02 |
|  | Benzodiazepines | 729 | 0.418<br>(0.318-0.550) | 4.18E-10 * | 2484 | 0.322<br>(0.232-0.447) | 1.48E-11 * | 611 | 1.02<br>(0.672-1.55) | 0.926 |

